# Identification and Mitigation of High-Risk Pregnancy with the Community Maternal Danger Score Mobile Application in Gboko, Nigeria

**DOI:** 10.1101/2021.08.18.21262256

**Authors:** Rajan Bola, Fanan Ujoh, Ronald Lett

**Affiliations:** Canadian Network for International Surgery, Vancouver, CANADA; Center for Sustainability and Resilient Infrastructure and Communities, London South Bank University, UNITED KINGDOM

## Abstract

**Introduction:** Nigeria constitutes 1% of the world’s population yet accounts for 10% of global maternal mortality. Risk analyses within rural regions of Nigeria are not routinely conducted, yet could help inform access to skilled birth care. The objectives of this study were to assess the proportion of women at risk for mortality or morbidity in Benue State, Nigeria by analysing data collected during routine antenatal visits and through the Community Maternal Danger Score (CMDS), a validated risk-analysis tool.

**Methods:** Two cohorts, comprised of pregnant women presenting to primary healthcare centres within Gboko, Benue State between 2015-2017 and 2020-2021, were included in this study. The 2015-2017 cohort had their risk assessed retrospectively through analysis of routinely collected data. Identification of risk was based on their age, parity, and disease status (HIV and diabetes). The 2020-2021 cohort had their risk assessed prospectively using the CMDS.

**Results:** Routinely collected data from 2015-2017 demonstrated that up to 14.9% of women in Gboko were at risk for mortality or morbidity. The CMDS reported that up to 21.5% of women were at a similar level of risk; a significant difference of 6.6% (p=0.006). The CMDS was more efficient in obtaining and assessing this data, and the identification was available in real-time for midwives and pregnant women.

**Conclusion:** Routine data collected in Gboko identifies a high proportion of pregnant women at risk for mortality or morbidity. The CMDS is an evidence-based risk analysis tool that expands on this assessment by also estimating individual and community-level risk, which allows for more efficient mitigation and prevention strategies of maternal mortality.

## Introduction

Records from the World Health Organization (WHO) indicate that sub-Saharan Africa accounts for 66% of global maternal deaths, with a maternal mortality ratio (MMR) of 546 per 100,000 live births.^1^ The Nigerian MMR is 512 per 100,000 live births,^2^ while we report an MMR for Benue State, one of Nigeria’s 36 states, of 1,189 per 100,000 live births.^3^ The National Demographic and Health Survey reveals that Nigeria constitutes approximately 1% of the world population yet accounts for 10% of the world’s maternal mortality rates.^2^ This trend has not improved: as of 2017 a Nigerian woman’s chance of dying from pregnancy and childbirth is 1 in 13, whereas it is 1 in 5,000 in developed nations.^4^ The United Nation’s Sustainable Development Goal Number 3 aims for a reduction of MMR below 70 per 100,000 live births.^5^ Countries like Nigeria would need to make considerable progress before achieving this goal.

Access to skilled birth attendants (SBAs) is a necessary requirement in maternal healthcare as it holds the potential to reduce maternal and infant mortality and morbidity rates in regions where they are employed.^6^ For developing countries, there is a proposed target of one SBA for every 5,000 population or one SBA to attend 200 births annually,^6-7^ while a lower target was set for SBAs in advanced and high-resource countries to attend between 30–120 deliveries annually.^8^ An SBA is a health professional, such as a midwife, doctor, or nurse who has been educated and trained to manage pregnancies, childbirth, and the immediate postnatal period.^9^ Midwives and nurses make up the majority of SBAs, yet have the least amount of formal training.^10^

SBAs also assist in the identification and management of complications in women and newborns, in addition to identifying abnormalities, managing them, or making referrals. They provide counselling on postnatal contraception to the mothers and actively assist in the prevention of mother-to-child transmission of HIV infection and other prevalent diseases.^11^ Studies show that countries have successfully reduced maternal mortality by focussing on training, recruiting, and supporting SBAs at deliveries.^1,12-14^ SBAs play a vital role in monitoring and counselling pregnant women on diseases such as HIV, diabetes, and syphilis,^11,15-17^ which have been noted for their high prevalence throughout low-to-middle income countries, including Nigeria.^18-20^

Several Nigerian studies have revealed variation in access to and utilisation of SBAs. Regional surveys indicate that only one-third of births take place in health facilities with SBAs, with the lowest recorded in North-west Zone (12%) and highest in the South-east Zone (78%).^21^ Urban areas such as Makurdi and Nnewi have reported over 80% of childbirths attended by SBAs. Estimates by the WHO suggest that approximately 59% of births in Nigeria were assisted by SBAs in urban areas, compared to 27% in rural areas.^22^ Traditional birth attendants are also commonly involved in birth practices in Nigeria, assisting in 13.5% of the pregnancies in Sagamu and 4.8% in Kaduna.^23-24^ Pregnant women are encouraged to travel to primary healthcare centres (PHCs) to deliver with an SBA, however, the physical distribution of PHCs has a major role in access; there are inequalities in the provision of PHCs across Benue State, which may be a contributory factor impeding access to SBAs by pregnant women.^25^

Analysis of routinely collected maternal data can identify pregnant women who are at risk of mortality or morbidity.^3^ The Community Maternal Danger Score (CMDS) is a low-cost, evidence-based maternal risk analysis tool that was validated in Makurdi, Nigeria to predict the need for skilled birth attendance and identify high risk for maternal death.^3^ The CMDS is designed to help pregnant women make informed decisions about where they should deliver, and helps midwives identify women at risk. The CMDS has been developed into a scoring tool that is available as an Android application to encourage pregnant women to deliver with SBAs. Pregnant women are assessed by healthcare workers in PHCs who use this scoring system at their initial visit to obtain a prenatal score out of 5 and at the 3^rd^ trimester to obtain a perinatal score also out of 5. The total score out of 10 points provides the midwives and the pregnant women with a quantitative measurement of their risk. A CMDS score of 3 out of 10 or higher is suggested to be the threshold of high risk for pregnant women.^3^

To strengthen the provision of risk information, women are provided their risk score in writing through SMS messages, while also advising them on ongoing antenatal care, being proactive about danger signs of their pregnancy, and other advice on seeking SBAs at PHCs and hospitals. This is done to encourage SBA-seeking behaviours prior to delivery, ultimately to reduce the incidence of maternal mortality in regions where it is implemented. The CMDS is based on 7 risk factors, however due to data collection in Benue State, only 3 of the 7 factors were noted to be routinely recorded in obstetrical assessments.^26^ These risk factors include age,^27^ parity,^28^ and co-existing medical conditions (human immunodeficiency virus (HIV) and diabetes).^29-30^

## Objectives

In this study, we evaluated the proportion of pregnant women at risk for mortality or morbidity using their age, parity, and disease status in one of the most populous local government areas (LGAs) of Benue State: Gboko LGA. Our initial objective was to assess the proportion of women at risk using available data from PHCs. Our second objective was to conduct a proof of concept using the CMDS app to illustrate how mobile health technologies like the CMDS can be used to help midwives identify women at risk for mortality or morbidity and improve maternal care by promoting skilled birth attendance.

## Methods

### Study Design and Setting

This is a cohort study of pregnant women presenting for antenatal care at PHCs within Gboko LGA. Benue State, located in North-central Nigeria, contains 23 LGAs, including the study location of Gboko (Figure 1). Benue state has a population of 5.7 million, which is approximately 3% of Nigeria’s total population. Gboko accounts for 7.6% of the State’s total population of 5.7 million. Gboko is the traditional seat of authority of the *Tiv*, who are the largest ethnic group in the State. Antenatal data in Gboko was collected by randomly sampling 50% of PHCs within the LGA (Figure 1). The population of interest for this study is women who presented for antenatal care in Gboko between 2015-2017 and 2020-2021. No exclusionary factors were imposed.

**Figure 1.**
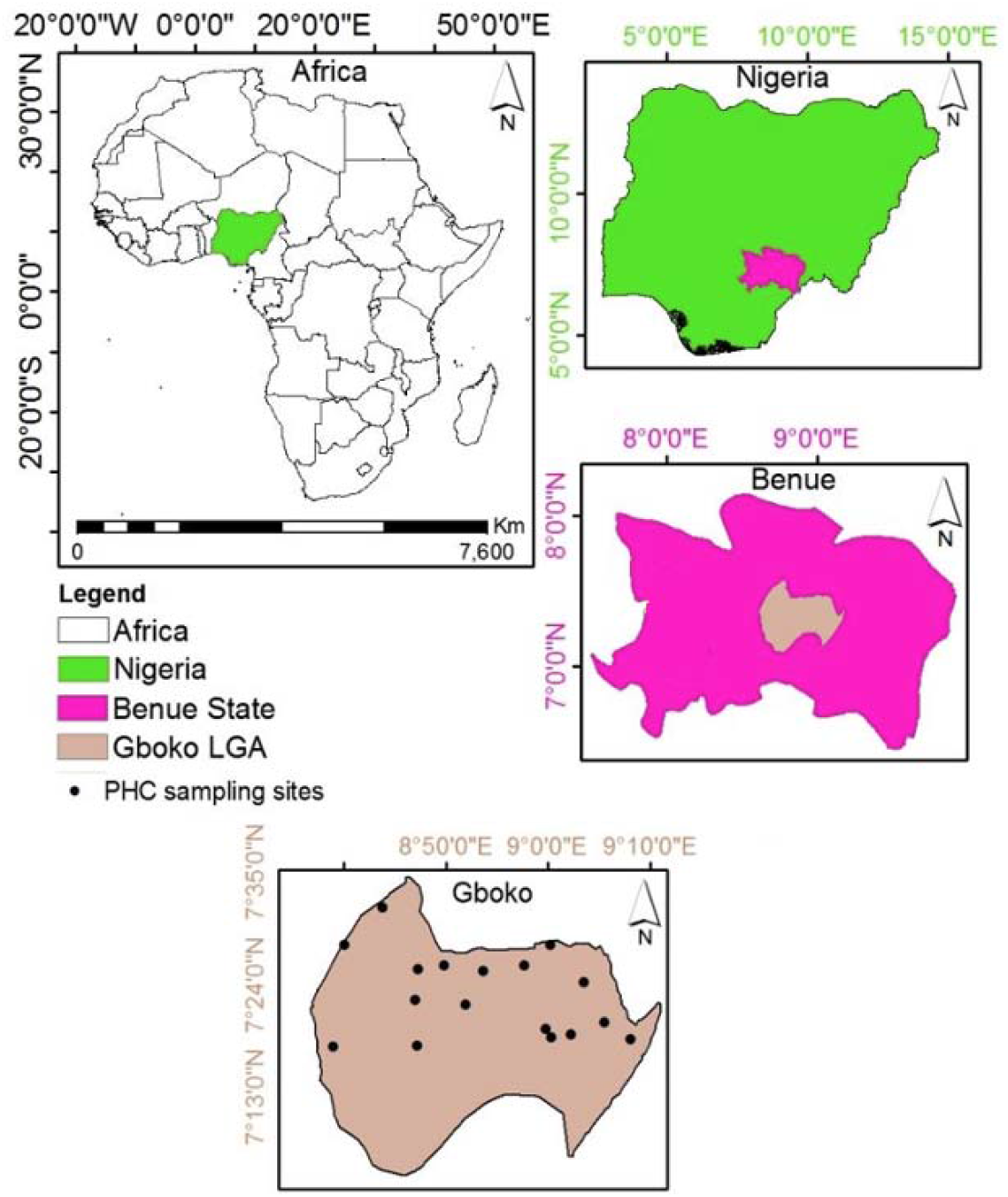
The study area of Gboko within the context of Benue State and Nigeria.

### Data Sources

Data from pregnant women over 3 years (2015-2017) was collected from existing records of 17 PHCs in Gboko in 2019. The proportion of women with risk factors was retrospectively ascertained using their age, parity, and disease status (HIV or diabetes). This was considered a routine risk identification through the available maternal data. Women from this retrospectively assessed cohort with up to 1 matching risk factor were considered *low risk*, women with 2 matching factors were considered *high risk*, and women with 3 factors were considered *extremely high risk*.^27-30^ The absolute proportion of women at risk was also assessed. All PHC data was categorized using definitions incorporated by the CMDS. Age was sorted into three categories: teens (ages 12-19), adults (ages 20-34), and older adults (ages 35 and older). The women were considered nulliparous if it was their first pregnancy, multiparous if pregnant between 1-4 times, and grand-multiparous if pregnant 5 or more times.

In addition, we prospectively applied the CMDS to a second cohort of women who presented to PHCs in Gboko between 2020-2021. Data collection included the full set of CMDS variables, and the women were scored according to the CMDS algorithm (Table 1). The proportion of women at risk were assessed, and their data was stored on a locally hosted online server which were exported as Excel files for analysis.

**Table 1.** Scoring Criteria of the CMDS for Women in Gboko (2020-2021).

| **Name of Risk Factor** | **Requirement for 2 Points** | **Requirement for 1 Point** | **Requirement for 0 Points** |
| --- | --- | --- | --- |
| Age | – N/A | – Below 20 years of age<br>– 35 years of age or older | – 20 years of age or older and below 35 years of age |
| Parity | – N/A | – Nulliparous (para = 1)<br>– Grand-multiparous (para > 4) | – Multiparous (para 1-4) |
| Patient Size | – N/A | - Underweight (BMI ≤ 18.5 kg/m<sup>2</sup>)<br>– Overweight (BMI > 30.0 kg/m<sup>2</sup>) | - Normal weight (BMI > 18.5 and ≤ 30.0 kg/m<sup>2</sup>) |
| Obstetrical History | Two of the following:<br>– Previous hemorrhage<br>– Previous stillbirth or miscarriage<br>– Previous breech delivery<br>– Previous twins (or more)<br>– Previous pregnancy within 1.5 years or greater than 5 years ago<br>– Reported reduction in fetal movements | One of the following:<br>– Previous hemorrhage<br>– Previous stillbirth or miscarriage<br>– Previous breech delivery<br>– Previous twins (or more)<br>– Previous pregnancy within 1.5 years or greater than 5 years ago<br>– Reported reduction in fetal movements | – Lack of these conditions |
| Fundal Height (3<sup>rd</sup> trimester) | – N/A | - Fundal height ≤35cm<br>– Fundal height >40cm | - Fundal height >35cm and ≤40cm |
| Signs of Pre-Eclampsia | - Very High Blood pressure (≥140 systolic and ≥100 diastolic mmHg) | – High blood pressure (>120 systolic or >90 diastolic mmHg) | - Normal blood pressure (≤120 systolic and ≤90 diastolic) |
|  | OR<br>Any sign of pre-eclampsia: proteinuria, headache, epigastric pain, blurred vision, excessive weight gain of 1kg or greater per week, or seizures<br>AND<br>High blood pressure (>120 systolic or >90 diastolic mmHg) |  | mmHg)<br>Lack of the aforementioned signs |
| Co-existing Conditions | Two of the following:<br>HIV<br>Anemia<br>Tuberculosis<br>Female genital mutilation<br>Diabetes<br>Malaria<br>OR<br>One of the following:<br>Maternal sepsis<br>Fever with ruptured membranes | One of the following:<br>HIV<br>Anemia<br>Tuberculosis<br>Female genital mutilation<br>Diabetes<br>Malaria | Lack of the aforementioned conditions |

### Statistical Analysis

Data analyses were based on basic descriptive statistics and Chi Square Test for proportions. Univariate analyses highlighting proportions and counts were calculated for age, disease status, and parity of all pregnant women. The proportion of women at risk for mortality or morbidity based on their age, parity, and disease status was estimated for the cohort of women from 2015-2017. We describe the median CMDS score, report interquartile ranges, and estimate the proportion of women at risk for the 2020-2021 cohort. We compared the proportions of women at risk for mortality or morbidity between the two cohorts from 2015-2017 and 2020-2021 using Chi Square Test. Statistical significance was assumed at p-value less than or equal to 0.05. RStudio was used for statistical analysis.

### Ethical Consideration

Ethical approval was sought and obtained from the Benue State Ministry of Health and Human Services, Makurdi granting access to antenatal data at PHCs in Gboko LGA. Part of the conditions listed in the ethical code is to ensure anonymity of the patients. Hence, personal identifiers are excluded in the distribution of the study outcome.

## Results

### Univariate Summary for Age, Parity, and Disease Status

The data collected from the retrospective cohort came from 17 PHCs and include a total of 604 patients (Table 2). Teens accounted for 14.2% of the sample, adults comprised 79.8%, while older adults made up 6.0%. Nulliparous women comprised 9.3% of the sample, multiparous women accounted for 81.4%, and grand-multiparous women accounted for 9.3%. 256 women had either diabetes or HIV, while 1 woman had both. 347 women did not have either HIV or diabetes.

**Table 2.** Counts and proportions of pregnant women in Gboko based on age, parity, and disease status measurements by cohort year.

| Condition and Category |  | 2015-2017 |  | 2020-2021 |  | p-value |
| --- | --- | --- | --- | --- | --- | --- |
|  |  | Count | Proportion | Count | Proportion | *marks a statistically significant difference |
| *Age* | Teenager | n=86 | 14.2% | n=72 | 16.6% | p=0.6773 |
|  | Adult | n=482 | 79.8% | n=320 | 73.9% | p=0.0504 |
|  | Older Adult | n=36 | 6.0% | n=41 | 9.5% | p=0.5717 |
| *Parity* | Nulliparous | n=56 | 9.3% | n=196 | 45.3% | p<0.0001* |
|  | Multiparous | n=492 | 81.4% | n=176 | 40.6% | p<0.0001* |
|  | Grand-Multiparous | n=56 | 9.3% | n=61 | 14.1% | p=0.4235 |
| *Disease Status* | No Diseases | n=347 | 57.5% | n=430 | 99.3% | p<0.0001* |
|  | HIV or Diabetes | n=256 | 42.3% | n=3 | 0.7% | p=0.1472 |
|  | HIV and Diabetes | n=1 | 0.2% | n=0 | 0% | N/A |

The data collected prospectively between 2020-2021 cover 433 patients (Table 2). Teens comprised 16.6% of the cohort, adults 73.9%, and older adults 9.5%. Nulliparous women accounted for 45.3% of the sample, while multiparous women accounted for 40.6%, and grand-multiparous accounted for 14.1%. Only 3 women were reported to have either diabetes or HIV.

### Routine Risk Identification and CMDS Comparison in Gboko

Of the 604 women from the retrospective cohort, 212 women were not identified to be at risk based on these factors, 302 women had 1 matching risk factor, 80 women had 2 risk factors, and 10 women had 3 risk factors. Up to 85.1% of women were at low risk for maternal mortality or morbidity, 13.2% were at high-risk for mortality or morbidity, and 1.7% were at extremely high risk for mortality or morbidity. At least 14.9% of women from the retrospective cohort would have needed an SBA to prepare for delivery.

The women who were scored prospectively using the CMDS guidelines in Table 1 (2020-2021) had a median score of 2 out of 10 with an interquartile range between 1 and 3. None of the women were scored as 0 out of 10. 191 women (44.1%) were scored as 1, 149 women (34.5%) were scored as 2, 72 women (16.6%) were scored as 3, 20 women (4.6%) were scored as 4, and 1 woman (0.2%) was scored as 5. The CMDS score distribution for this cohort of women is shown in Figure 2. Based on the CMDS threshold of risk of 3 points or higher, 21.5% of women from the prospective cohort were provided SMS text messages and warning messages to encourage them to seek an SBA in preparation for delivery.

**Figure 2.**
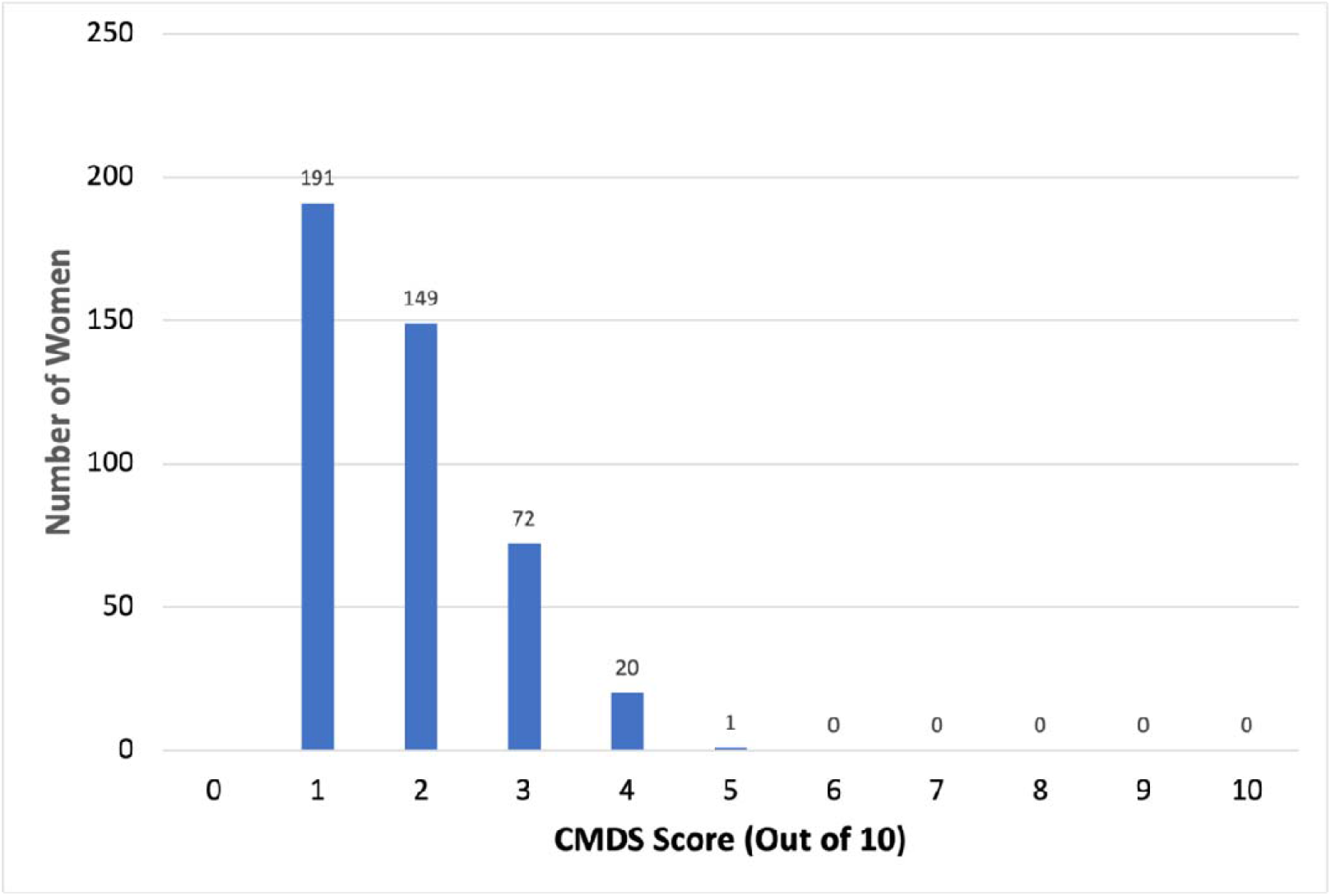
Distribution of CMDS Scores for a cohort of pregnant women in Gboko, 2020-2021.

Based on the routine risk identification for the 2015-2017 cohort of women, which identified 14.9% of women at risk, and CMDS scoring for the 2020-2021 cohort of women, which identified 21.5% of women at risk, there is a significant difference between these proportions of 6.6% (p=0.006). Thus, the CMDS was not only able to identify more women at high risk than routine data collection, but was also able to do this in real-time.

## Discussion

For the first time in the literature, an estimate of the proportion of women at risk for maternal mortality or morbidity in Benue State has been reported. The inclusion of two temporally distinct cohorts in this study allowed for the determination of maternal characteristics of pregnant women in Gboko (Table 2). It was found that the general constitution of the population has not considerably changed in the prior 4 years, however there are slight differences in the proportions of women seeking skilled birth care based on their parity (p<0.0001) and disease status (p<0.0001).

The routine risk analysis in Gboko demonstrated that between 2015-2017, approximately 15% of women needed an SBA in preparation for a safe delivery. This result is the first analysis of risk in Benue State depicting a large proportion of pregnant women who require an SBA at delivery. Local healthcare teams and policy makers do not routinely collect maternal data to assess baseline risk in the community. However, these findings demonstrate that there is a great proportion of women at risk within the community, and that local decision makers should be monitoring these community-level characteristics to allocate lifesaving resources to prevent this phenomenon from adversely impacting the MMR of Benue State.

The CMDS was used to score the cohort of pregnant women from 2020-2021 to ascertain the level of risk using a more comprehensive assessment developed from evidence-based maternal risk factors. In addition to reporting a significantly higher proportion of women at risk of 21.5% (p=0.006), the CMDS was also able to identify this level of risk in real-time, and subsequently recommend prevention strategies. This is relevant given that midwives and community healthcare workers in rural areas are expected to care for a large number of pregnant women at high risk, yet often lack the tools required to conduct risk assessments to inform their needs and the women’s necessity to access SBAs within Gboko.

Benue State has the potential to decrease maternal mortality by identifying women at high risk and mitigating that risk. By seeking skilled care, the women are more likely to experience a positive birthing outcome. Thus, more SBAs, adequate distribution of obstetrical resources, and improved antenatal data sources are required to support the needs of pregnant women at risk in Benue State. Without methods to identify these women, and infrastructure to manage this risk, the MMR in Benue State will likely remain unchanged. This study supported the need for improved antenatal data collection for other important demographic and maternal health variables.^18^ Improved data collection through the use of mobile health applications like the CMDS would allow for standardized and comprehensive data which could support policy and decision making.^30^

The CMDS was designed to improve the maternal health outcomes of women in low-resource settings where accessing skilled care can be burdensome and where routine data is only available years later and inaccessible to the women studied. In this study, the CMDS was shown to identify women at risk for mortality in a manner that was more efficient than conventional data management methods at PHCs. The CMDS provided comprehensive assessment on an expanded set of variables to standardize and improve on data collected at PHCs, while also incorporating best-practice guidelines.^3^ Integrating the CMDS in routine data collection efforts would prove to be an effective strategy for collecting maternal data across rural regions. This data can then be used to inform health resources planning, epidemiological evaluations, and risk assessments within Gboko and other regions impacted by high MMR. By implementing the CMDS, the care of pregnant women can be improved such that the MMR can be reduced to manageable targets, like those proposed within the Sustainable Development Goals.^5^

## Limitations

The primary limitations of this study were that the 2015-2017 cohort depended on existing antenatal records, while the 2020-2021 cohort was limited in sample size. In addition, both cohorts only included data from women who were seeking care at PHCs. Due to limited time and resources, it was not feasible to collect data for women who did not seek care at PHCs. The CMDS may benefit community extension workers who are in contact with, thus future studies should collect data on a sample of women who did not seek skilled care at PHCs.

## Conclusion

We demonstrate that the current antenatal data collection efforts at primary healthcare centres are insufficient to support community-level risk assessments within the State due to the limited number of variables that are routinely collected. Mobile health technologies like the Community Maternal Danger Score can efficiently standardize data collection and inform healthcare planning and resource allocation, ultimately to improve maternal care. Future studies should assess the impact on mobilization and improvements to maternal outcomes that the Community Maternal Danger Score has in low-resource settings.

## Data Availability

The datasets used and/or analysed during the current study are available from the corresponding author on reasonable request.

## Supplemental Information

### Availability of data and materials

- The datasets used and/or analysed during the current study are available from the corresponding author on reasonable request.

### Funding

- Funding was provided by Grand Challenges Canada. The grant was awarded directly to the authors’ institution: the Canadian Network for International Surgery. Grand Challenges Canada receives support through Global Affairs Canada. The website for the Grand Challenges Canada can be found at: https://www.grandchallenges.ca.

## Acknowledgements

- We would like to acknowledge Paul Aganyi of the Federal Medical Centre, Elizabeth Akpehe of the Health Department in Gboko LGA, and Levi Kwaghngee of the Health Department in Gboko LGA for their contributions and assistance with developing and operating the CMDS Application in Nigeria.

## Author’s Contributions

- FU, RB, and RL conceived of the hypothesis. FU verified the theory, collected data, and collaborated with local stakeholders to access local perinatal records. RL supervised the project and contributed to the interpretation of the results. RB and FU prepared the manuscript with assistance from RL.

